# Diagnostic Reversion in Dementia Care: A Real-World Analysis of Mild Cognitive Impairment Diagnoses Following Dementia in a Large Electronic Medical Record System

**DOI:** 10.1101/2025.07.16.25331678

**Authors:** Silvia Miramontes, Umair Khan, Scott Zimmerman, Erin L. Ferguson, Hunter Mills, Boris Oskotsky, Evan Phelps, Tomiko T. Oskotsky, Anthony J. Capra, M.Maria Glymour, Marina Sirota, Elena Tsoy

## Abstract

**INTRODUCTION:** If mild cognitive impairment (MCI) is diagnosed after dementia, it suggests either the dementia diagnosis was premature, or the MCI diagnosis is incorrect. We investigated the prevalence and predictors of such “diagnostic reversion”—MCI diagnosis following dementia diagnosis—in a large academic health system.

**METHODS:** Among 5,965 patients aged 50+ with incident dementia in UCSF Health electronic health records, we identified “reverters” with a subsequent MCI diagnosis. We used Group LASSO–regularized logistic regression and random forest models to identify predictors.

**RESULTS:** Reversion occurred in 13.7% of patients. Lower odds were observed among older adults (OR=0.95/year; 95% CI: 0.92–0.98), while higher odds were found among Spanish speakers (OR=2.26; 95% CI: 1.28–4.00), those with greater cardiovascular risk (OR=1.16; 95% CI: 1.01–1.33), and higher Charlson comorbidity burden (OR=1.09; 95% CI: 1.05–1.14).

**DISCUSSION:** Diagnostic reversion is common and socially patterned, suggesting contributions from misdiagnosis, clinical uncertainty, or variability in clinical presentation and care setting.

## 1. Background

Accurate diagnosis of cognitive impairment and dementia is essential for appropriate treatment, care planning, and identification of participants for clinical trials. Mild cognitive impairment (MCI) is often conceptualized as a transitional state between healthy cognitive aging and dementia [1], but the distinction between these conditions can be nebulous in real-world clinical practice. Clinical diagnostic criteria for dementia and MCI are well-established, but inconsistencies in the timing and accuracy of diagnosis may still occur in routine care [2–4].

Dementia is frequently under- or misdiagnosed in routine care settings [3,5]. In the Aging, Demographics, and Memory Study (ADAMS), fewer than 50% of participants who met diagnostic criteria for dementia during the research assessment had ever received a cognitive evaluation by a physician [6]. In primary care, cognitive impairment often goes unrecognized– one study found that over 40% of cognitively impaired patients were not identified by their providers [7]. A recent meta-analysis further estimated that 61% of dementia cases in the U.S. remain undiagnosed [8]. These diagnostic challenges are especially pronounced outside of specialty or memory clinics, where clinicians often rely on brief cognitive screening tools or subjective impressions rather than comprehensive assessments [4,9]. In these settings, time constraints, limited access to specialists, and variability in how cognitive symptoms present can hinder diagnostic accuracy and delay recognition [10]. Social and structural factors further compound these limitations. For example, patients with lower educational attainment, who live alone, come from low-income backgrounds, or have poorer health are more likely to experience delayed or missed diagnoses [11–15]. Such disparities reflect broader issues in diagnostic equity, as marginalized groups often face systemic barriers to timely and accurate dementia care.

Despite growing recognition of the challenges in diagnosing cognitive impairment [16–18], little is known about the stability or consistency of dementia diagnoses once assigned in routine clinical care [19–22]. Most prior work has focused on barriers to initial detection at the MCI stage [23], but less attention has been paid to potential diagnostic reversals or ICD code reclassifications following a dementia diagnosis. This represents a critical gap, as inconsistent diagnoses may reflect variability in clinical judgement, access to follow-up care, or patient communication, and could have profound implications for treatment, care planning, and patient autonomy [5]. Electronic health record (EHR) data provide a unique opportunity to investigate these patterns at scale by using real-world patient cohorts. In this study, we examine the frequency of diagnostic reversals from dementia to MCI and their associations with care setting factors and patient characteristics within a large academic EHR system.

## 2. METHODS

### 2.1 Study Population and Data Sources

We used electronic health record (EHR) data from the University of California, San Francisco (UCSF) Health system to identify patients aged 50 years and older who received a new (index) diagnosis of dementia between 1988 and 2024. We assessed diagnostic trajectories by classifying patients into two mutually exclusive groups: (1) the reference group, consisting of patients diagnosed with dementia who did not subsequently receive an MCI diagnosis, and (2) the reversion group, defined as patients who were initially diagnosed with dementia but who subsequently received an MCI diagnosis (**Fig. 1A**). Diagnostic codes for MCI and dementia were identified using the International Classification of Diseases, Ninth and Tenth Edition (ICD-9 and ICD-10) (list of codes provided in **Supplemental Tables S1 and S2**). We also included all ICD codes recorded prior to index dementia diagnosis to capture each patient’s comorbidity history.

**Figure 1:**
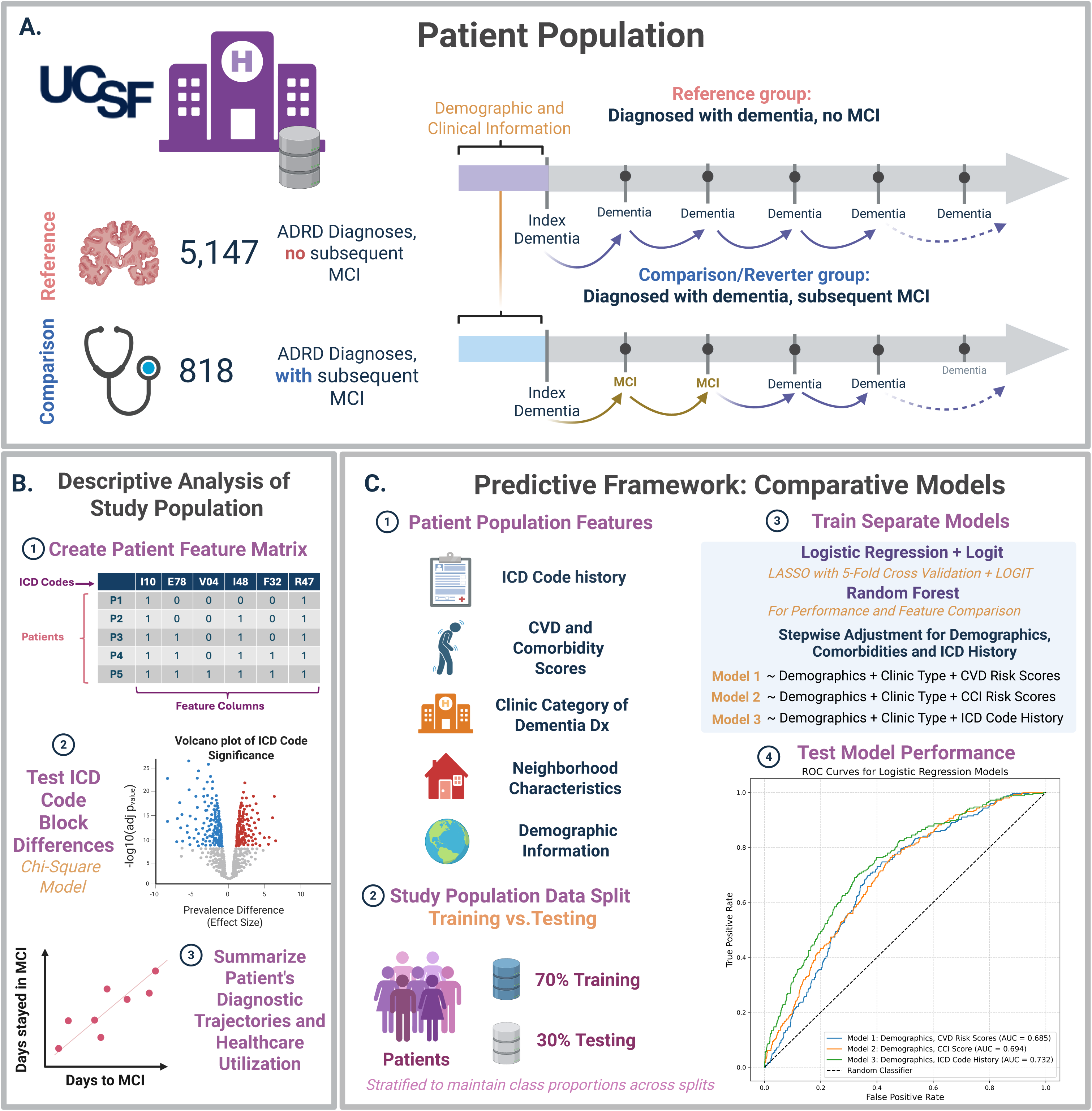
Study Design Schematic Illustrating Patient Population, Descriptive Analysis, and Predictive Modeling Framework. **A.** Patient population: Patients were identified from UCSF Health records based on an index diagnosis of dementia. The reference group (n = 5,467) included patients who never received a subsequent diagnosis of mild cognitive impairment (MCI). The comparison group (n = 818) included patients who were diagnosed with MCI after their initial dementia diagnosis, reflecting potential diagnostic reversion. **B.** Descriptive analysis of study population: (1) A patient-level feature matrix was constructed using all ICD code blocks recorded prior to the index dementia diagnosis. (2) Differences in ICD block code prevalence between patient groups were tested using a chi-square model, and results were visualized using a volcano plot. (3) Diagnostic trajectories were summarized by examining days to first MCI diagnosis and duration of time spent in the MCI state. **C.** Predictive framework: (1) Features included demographics, ICD code-block history, clinic category of dementia diagnosis, cardiovascular and comorbidity scores, and neighborhood characteristics. (2) The population was split into 70% training and 30% testing sets, stratified to maintain class proportions within splits. (3) Logistic regression (with LASSO-based feature selection followed by LOGIT) and random forest models were trained separately across three specifications, with adjustment for clinical demographic features. (4) Model Performance was evaluated and compared using ROC curves on the test set.

We extracted demographic variables including age at the time of index dementia, sex (reference: female), race/ethnicity (reference: White), and preferred spoken language (reference: English). The specialty of the clinic where the index dementia diagnosis was assigned was categorized into four clinic types: dementia clinics (including memory and geriatrics, as the reference category), primary care/internal medicine, emergency department, and other specialty clinics (e.g., Diabetes, Cardiology, Oncology; full list of categorized clinics as “other specialty clinics” in **Supplemental Table S3**).

### 2.2 Descriptive Analyses

#### 2.2.1 ICD-Based Feature Matrix and Chi-Square Comparisons

To construct a patient-level feature matrix, we derived comorbidity variables using structured ICD-9 and ICD-10 codes for all conditions ever recorded in the EHR prior to index dementia diagnosis. This approach captured the full history of comorbidities leading up to the index date, enabling us to characterize each patient’s baseline clinical profile, defined as the patient’s cumulative comorbidity burden prior to receiving the first dementia diagnosis. We selected this timeframe to reflect pre-existing health status, before any diagnostic reversion (e.g., dementia to MCI) could occur.

To facilitate interpretability, we grouped related ICD-9 and ICD-10 codes into higher-level *ICD block codes* to reflect clinically meaningful diagnostic categories (e.g., I67.2: Cerebral aneurysm and I67.2: Cerebral atherosclerosis are grouped under I67: Other cerebrovascular diseases). We then summarized each patient’s ICD block code history into a binary feature matrix (presence/absence of each ICD block code). To identify diagnostic block code differences between the two patient groups, we conducted chi-square tests with multiple hypothesis correction (Benjamini-Hochberg, significance threshold of p<0.01). ICD block codes present in fewer than 5% of the sample were excluded to reduce sparsity and improve interpretability, reducing the number of unique block codes from 1,931 to 101 (**Fig. 1B**).

#### 2.2.2 Characterizing Diagnostic Reclassification and Healthcare Utilization

Among patients in the reversion group, we summarized diagnostic reversal trajectories, including: (1) number of days from index dementia diagnosis to first MCI reversion; (2) number of days spent with an MCI diagnosis before further diagnostic changes (e.g., progression to a second dementia diagnosis); and (3) the proportion of patients who progressed to a second dementia diagnosis after MCI reversion. We also fit a linear regression model to evaluate whether time to first MCI reversion was associated with time spent with the reclassified MCI diagnosis (log-transformed for normality). Additionally, we compared the extent and duration of health system utilization across patient groups. We calculated (1) total days of pre-index observation time in EHR; (2) total number of ICD codes recorded; (3) number of unique ICD code blocks, representing distinct condition categories; and (4) number of unique healthcare visit dates captured in the EHR. Pre-index metrics were restricted to diagnoses and visits occurring before index dementia diagnosis, while total metrics included the full EHR history available per patient (**Fig. 1B**).

### 2.3 Covariate Construction

#### 2.3.1 Comorbidity Indices

To characterize each patient’s clinical profile, we constructed three continuous comorbidity indices using all ICD block codes recorded prior to index dementia diagnosis. First, we calculated a Cardiovascular Disease (CVD) Burden Score, defined as the count of diagnosed cardiovascular diseases (e.g., atrial fibrillation, heart failure), harmonized with the National Alzheimer’s Coordinating Center’s Uniform Data Set Participant Health History and Clinician-assessed Medical Conditions Form [24]. Second, we constructed a Cardiovascular (CV) Risk Score, reflecting the number of common cardiovascular risk factors (e.g., hypertension, diabetes, hyperlipidemia) [25], excluding conditions already accounted for in the CVD Burden Score. Lastly, we calculated the Charlson Comorbidity Index (CCI) using the R ‘comorbidity’ package by mapping patient ICD codes to comorbidity conditions based on established algorithms [26,27]. Full ICD code groupings used for each CVD-related score are provided in **Supplemental Tables S4 and S5**.

#### 2.3.2 Socioeconomic Variables

To assess the role of neighborhood socioeconomic context in diagnostic reversion, we linked each patient’s residential census tract (as recorded on or before index dementia diagnosis) to data from the U.S. Census and American Community Survey [28] via the UCSF Health Atlas [29,30]. This geographic unit was treated as a baseline exposure, reflecting patients’ neighborhood context at the time of index dementia diagnosis. We extracted census tract-level educational attainment using three variables: the proportions of residents with (a) less than a high school diploma; (b) a high school diploma or more; and (c) a bachelor’s degree or higher. All education percentiles were converted to deciles to aid model interpretability. Additionally, we included median home value (scaled per $100,000 USD) as a continuous measure of neighborhood socioeconomic status.

### 2.4 Predictive Framework

We implemented a comparative modeling framework to evaluate how different algorithms perform when predicting diagnostic reversion to MCI. Specifically, we trained both logistic regression and random forest classifiers to identify patient-level characteristics and comorbidities associated with the likelihood of diagnostic reversion to MCI following an index dementia diagnosis.

To assess model generalizability and performance, we randomly split the study population into a training set (70%) and a test set (30%), using stratified sampling based on the binary outcome to preserve class proportions. All model fitting and feature selection were conducted on the training set, with final performance and coefficient interpretation evaluated on the test set (**Fig 1C**).

#### 2.4.1 Logistic Regression

We implemented a two-step logistic regression approach to identify and evaluate the most predictive features associated with diagnostic reversion to MCI, using three conceptually distinct comorbidity variable sets across models (**Fig 1C**). First, we applied Group LASSO-regularized logistic regression with 5-fold cross-validation on the training set, using a grid search to tune the group-level regularization strength. Specifically, we varied the group regularization parameter C (the inverse of regularization strength λ), across a log-scaled range from 1e-6 to 1e3. Group-level penalization shrank entire sets of related features (e.g., one-hot encoded categories), enabling the model to retain only the most predictive groups while discarding those that contributed little to classification.

We evaluated Group LASSO and LOGIT models using three different covariate sets (**Fig. 1C**):

1. Demographics + clinic type + CVD Burden + CV Risk Scores
2. Demographics + clinic type + Charlson Comorbidity Index (CCI)
3. Demographics + clinic type + ICD block-level codes recorded prior to index dementia diagnosis

Baseline demographic features included age, sex, preferred spoken language, median home value (scaled by $100,000 USD), and three tract-level education variables (converted to deciles). Categorical variables (preferred language and clinic type) were one-hot encoded, and continuous features were standardized prior to model fitting.

Predictors with non-zero coefficients from Group LASSO model were used to refit standard logistic regression (LOGIT) and estimate β coefficients with 95% confidence intervals. These LOGIT models were evaluated on the test set to assess generalizability and coefficient stability. By comparing effect sizes across the training and test sets using the same Group LASSO–selected predictors, we evaluated whether associations reflected reproducible signals rather than overfitting.

#### 2.4.2 Random Forest Models

To determine whether more complex, non-parametric models could improve predictive performance over the interpretable coefficients of the two-step logistic regression approach, we also implemented random forest models using the same three covariate configurations. While random forests do not produce coefficient estimates, they provide feature importance scores that summarize how often and effectively a feature is used to split the data in the trees that make up the model. Importantly, these scores capture a feature’s *influence* on model decisions but may be affected by feature correlations and are not measures of effect size. To focus on the most informative features while reducing noise from low-signal variables, we retained predictors that together explained 90% of the model’s cumulative importance, a data-driven threshold capturing the majority of the model’s predictive signal.

#### 2.4.3 Model Performance Comparison

We evaluated model discrimination using the Area Under the Receiver Operating Characteristic Curve (AUROC) [31] to assess how well each predictive approach distinguished patients likely to experience diagnostic reversion based on comorbidities and demographic characteristics. To address the impact of class imbalance (∼14% reversion cases in our data splits) [32,33], we also calculated the Area Under the Precision-Recall Curve (AUPRC) [34,35], which is more sensitive to performance on the minority class.

While AUROC and AUPRC provide valuable threshold-independent measures of discrimination, they do not indicate how a binary classifier would perform when assigning class labels using a specific probability cutoff. To address this, we implemented threshold tuning (also known as “threshold moving”): after training each model, we selected the probability threshold on the training set that maximized the F1 score (harmonic mean of precision and recall), based on the precision recall curve. This approach is increasingly recognized as a simple yet effective strategy for improving minority-class detection in imbalanced classification problems [36–39]. We applied the selected threshold to the test set to estimate classification performance, including sensitivity (true positive rate, TPR), specificity (true negative rate, TNR), precision (positive predictive value, PPV), and F1 score.

Together, these complementary metrics offer a comprehensive view of model utility: AUROC reflects overall ranking ability, AUPRC captures positive-class tradeoffs, and threshold-based metrics quantify real-world classification performance under both default and optimized probability cutoffs.

## 3. RESULTS

We analyzed EHR data from 5,965 patients with index dementia diagnoses aged 50 years and older. The sample included a reference group (N=5,147; 86.3%) who received a dementia diagnosis without a subsequent MCI diagnosis, and a reversion group (N=818; 13.7%) who were diagnosed with MCI following an initial dementia diagnosis. Demographic and clinical characteristics of both groups are summarized in **Table 1**.

**Table 1:** Baseline Characteristics of Patients (Reference and Comparison Groups)

|  | Reference:<br>Dementia Only<br>(N=5,148) | Comparison:<br>Reverters<br>(N=818) | P-value |
| --- | --- | --- | --- |
| **Age at Baseline (years)** |  |  |  |
| Mean (SD) | 74.7 (8.62) | 71.1 (6.87) | <0.001 |
| Median [Min, Max] | 75.0 [50.0, 90.0] | 71.0 [45.0, 87.0] |  |
| **Sex (Ref: Female)** |  |  |  |
| Female | 3125 (60.7%) | 508 (60.2%) | 0.694 |
| Male | 2022 (39.3%) | 310 (39.8%) |  |
| Unknown | 1 (0.0%) | 0 (0%) |  |
| **Race/Ethnicity (Ref: White)** |  |  |  |
| White | 2698 (52.4%) | 485 (59.3%) | <0.001 |
| Asian | 775 (15.1%) | 109 (13.3%) |  |
| Black or African American | 357 (6.9%) | 53 (6.5%) |  |
| Latinx | 353 (6.9%) | 74 (9.0%) |  |
| Other Identity | 499 (9.7%) | 82 (10.0%) |  |
| Unknown/Declined | 466 (9.1%) | 15 (1.8%) |  |
| **Preferred Language (Ref: English)** |  |  |  |
| English | 3825 (74.3%) | 645 (78.9%) | <0.001 |
| Chinese - Cantonese | 399 (7.8%) | 48 (5.9%) |  |
| Chinese - Mandarin | 102 (2.0%) | 13 (1.6%) |  |
| Other | 311 (6.0%) | 37 (4.5%) |  |
| Russian | 300 (5.8%) | 22 (2.7%) |  |
| Spanish | 211 (4.1%) | 53 (6.5%) |  |
| **Clinic Type (Ref: Dementia Spec.)** |  |  |  |
| Dementia Specialty | 1563 (30.4%) | 353 (43.2%) | <0.001 |
| ER | 137 (2.7%) | 8 (1.0%) |  |
| Other | 1716 (33.3%) | 115 (14.1%) |  |
| Primary Care or Internal Medicine | 1732 (33.6%) | 342 (41.8%) |  |
| **CVD Burden Score** |  |  |  |
| Mean (SD) | 0.58 (1.05) | 0.408 (0.89) | <0.001 |
| Median [Min, Max] | 0 [0, 7.00] | 0 [0, 6.00] |  |
| **CV Risk Factor Score** |  |  |  |
| Mean (SD) | 1.09 (1.17) | 1.09 (1.11) | 0.946 |
| Median [Min, Max] | 1.00 [0, 5.00] | 1.00 [0, 4.00] |  |
| **Charlson Comorbidity Index** |  |  |  |
| Mean (SD) | 2.65 (3.25) | 3.27 (3.56) | <0.001 |
| Median [Min, Max] | 2.00 [0, 22.0] | 2.00 [0, 24.0] |  |
| **% Bachelors or More (per 10%)** |  |  |  |
| Mean (SD) | 5.33 (2.02) | 5.69 (1.94) | <0.001 |
| Median [Min, Max] | 5.54 [0.10, 9.18] | 5.70 [0.32, 9.18] |  |
| **% < less than High School (per 10%)** |  |  |  |
| Mean (SD) | 1.02 (0.89) | 0.91 (0.82) | < 0.001 |
| Median [Min, Max] | 0.78 [0, 6.61] | 0.69 [0, 5.97] |  |
| **% High School or More (per 10%)** |  |  |  |
| Mean (SD) | 8.98 (0.89) | 9.09 (0.82) | < 0.001 |
| Median [Min, Max] | 9.22 [3.39, 10.0] | 9.31 [4.03, 10.0] |  |
| **Median Home Value (per \$100K USD)** | | | |
| Mean (SD) | 10.2 (4.41) | 10.9 (4.22) | < 0.001 |
| Median [Min, Max] | 10.00 [0.48, 20.0] | 10.7 [1.65, 20.0] |  |

Baseline characteristics of patients with dementia diagnoses with and without subsequent MCI diagnoses. P-values are derived from t-tests for continuous variables and chi-square tests for categorical variables. P-values assess differences between the reference group and the comparison group.

### 3.1 Baseline Differences in Reversion Timing, Comorbidity Profiles and Healthcare Utilization

Among the 818 patients who reverted to MCI after an initial dementia diagnosis, the median time to first reversion was 283 days (IQR: 95 – 837; range: 1 – 10,873 days). After reclassification, patients continued with their MCI diagnosis for a median of 90 days (IQR: 13 – 313; range: 0 – 5,132 days). Notably, 73.4% of reverters (N=600) later received a second dementia diagnosis, with a median time to reclassify to the second dementia diagnosis of 98 days (IQR: 32–310; range: 1 – 4,657 days). A log-log scatter plot revealed a weak but positive nonlinear association between time to MCI reversion and duration spent in MCI (**Fig. 2A**, **2B**). This trend was visualized using locally weighted scatterplot smoothing (LOWESS) line, fit separately for patients who reverted to MCI within one year and those who reverted after one year.

**Figure 2:**
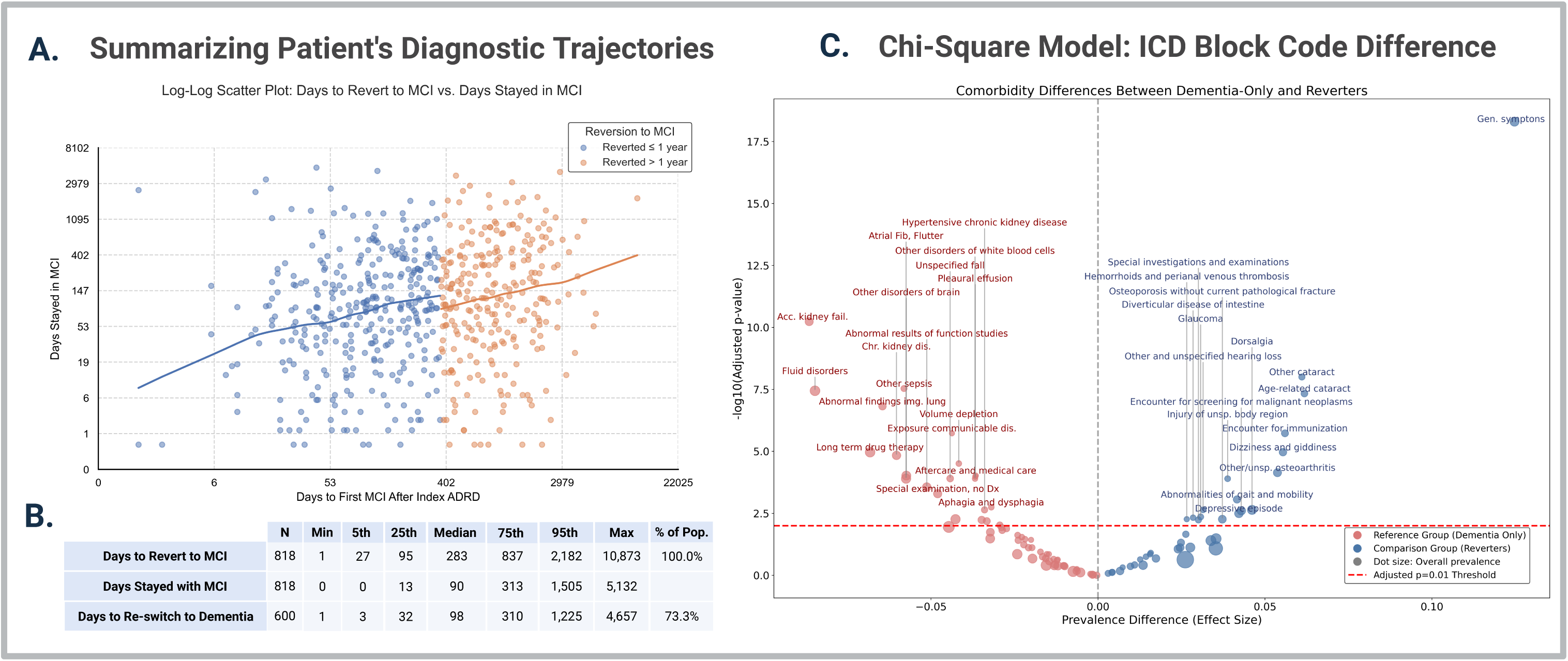
Descriptive Analysis of Study Population. **A.** Log-log scatter plot of time to reversion to MCI versus duration spent in the MCI state, among patients who reverted after an initial dementia diagnosis. The plot is stratified by timing of reversion: patients who reverted to MCI within one year of their index ADRD diagnosis (blue), and those who reverted after more than one year (orange). Axes are log-scaled to improve visibility across a wide range of follow-up duration. Locally weighted scatterplot smoothing (LOWESS) was applied to each subgroup to estimate the nonparametric regression lines. **B.** Summary statistics of reverter group. From the sub-population of N=818, we display the percentiles of the number days taken to revert to MCI after index Dementia diagnosis, the number of days continuing with MCI diagnosis, and the number of days taken to re-switch to a Dementia diagnosis. **C.** Chi-Square model: ICD block code differences. Volcano plot displaying differences in ICD block-level code prevalence between patients who reverted to MCI (Group 1) and those who remained diagnosed with dementia (Group 2). The x-axis shows the difference in prevalence (Group 1-Group 2), and the y-axis shows the -log10 of the Benjamini Hochberg-adjusted p-values from chi-square tests. Prevalence was defined as the proportion of patients in each group who had a given ICD block code recorded prior to the index dementia diagnosis. Points on right side represent code blocks more prevalent among patients who reverted to MCI; points on the left represent code blocks more prevalent among patients who remained diagnosed with dementia. Dot size corresponds to the overall prevalence of each ICD block code across the full sample. The dashed horizontal line indicates the significance threshold (p < 0.01 after correction).

We compared overall healthcare utilization between groups (**Supplemental Tables S6 — S7**). While pre-index observation time and visit frequency were similar, a larger proportion of Dementia Only patients had no documented diagnoses before their index date. Post-index, reverters had significantly longer follow-up, more recorded diagnoses, and more unique conditions and visit days, indicating greater overall EHR engagement.

Chi-Square analyses yielded 39 ICD block-level codes that were significantly different between the reference and reverter groups after multiple-hypothesis test correction (Benjamini-Hochberg, p < 0.01). Among these codes, those with higher prevalence in the reference group included chronic kidney disease (N18), atrial fibrillation (I48), fluid and electrolyte disorders (E87), abnormal findings on imaging of lung (R91), long-term drug therapy (Z79), disorders of the brain (G93), and respiratory disorders (J98). In contrast, codes more prevalent in the reverter group included dizziness and giddiness (R42), depressive episode (F32), osteoarthritis (M19), cataracts (H25/H26), dorsalgia (M54), and unspecified injury (T14) (**Fig. 2C**). A complete list of all ICD block codes identified in Chi-Square analyses can be found in **Supplemental Table S8**.

### 3.2 Comorbidities and Demographic Factors Associated with Reversion to MCI in Logistic Regression Models

In the model configuration including demographics, preferred language, clinic type, CVD Burden, and CV Risk Scores, LASSO selected 16 features from the training set, used to fit a standard logistic regression to evaluate coefficient stability in the held-out test set (**Fig. 3A**, **3B**). In the test set, Spanish-language preference (OR=1.96, 95% CI: 1.08–3.53) and higher CV Risk Score (OR=1.18, 95% CI: 1.03–1.36) were associated with significantly greater odds of reversion to MCI. In contrast, increasing age (OR=0.96, 95% CI: 0.94–0.98), index dementia diagnoses made in “other specialty clinics” (OR=0.34, 95% CI: 0.22–0.52), and Russian-language preference (OR=0.32, 95% CI: 0.12–0.85) were consistently associated with lower odds of MCI reversion. A higher CVD Burden Score was significantly associated with lower odds of MCI reversion in the training set; however, this association did not replicate in the test set (OR=0.95, 95% CI: 0.80–1.12). Similarly, patients initially diagnosed with dementia in emergency room (ER) settings had lower odds of MCI reversion (OR=0.30, 95% CI: 0.13 – 0.65), though this too was not significant in the test set. All effect estimates remained directionally consistent with training estimates, but confidence intervals in the test set were wider, likely reflecting reduced statistical power.

**Figure 3:**
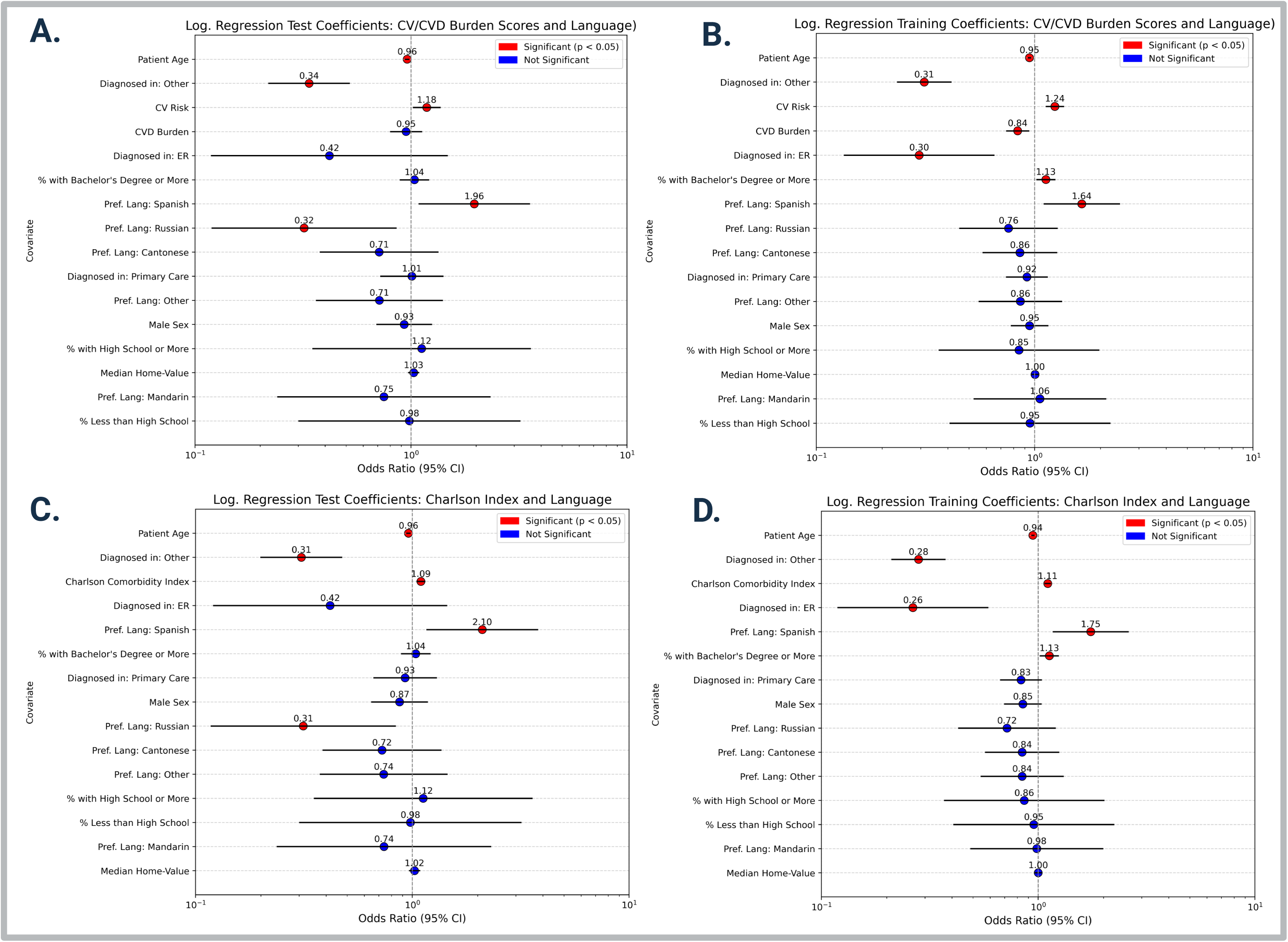
Logistic regression (LOGIT) coefficients from training and testing sets for models adjusted by preferred spoken language. All models include patient age at index dementia diagnosis, clinic type assigning the diagnosis, biological sex, and preferred language as covariates. **Panels A and B** present odds ratios (95% CI) for models including cardiovascular (CV) risk and cardiovascular disease (CVD) burden scores, fit on the test set (A) and training set (B), respectively. **Panels C and D** display coefficients from models including the Charlson Comorbidity Index as the primary clinical predictor, for the test set (C) and training set (D). Covariates shown represent the subset of non-zero predictors selected ruing the first step of a two-step logistic regression framework, in which group lasso with cross-validation was used for variable selection. Statistically significant predictors (p < 0.05) are shown in red; non-significant predictors are show in blue. Odds ratios are plotted on a log scale.

The second logistic regression configuration included demographics, preferred language, clinic type, and the Charlson Comorbidity Index (CCI). LASSO retained 15 predictors from the training set, fitted in a standard logistic regression to estimate effect sizes and assess generalizability in the held-out test set (**Fig. 3C**, **3D**). In the test set, higher odds of reversion to MCI were associated with Spanish-language preference (OR=2.10, 95% CI: 1.17–3.78), and higher CCI Scores (OR = 1.09, 95% CI: 1.05–1.14). Conversely, increasing age (OR=0.96, 95% CI: 0.94–0.98), index dementia diagnoses made in other specialty clinics (OR=0.31, 95% CI 0.20–0.47), and Russian-language preference (OR=0.31, 95% CI: 0.12–0.83) were significantly associated with lower odds of diagnosis reclassification to MCI. We found no significant association between census tract–level education or median home value and the risk of reversion to MCI in the test sets. However, in the training sets, a 10% increase in the proportion of neighborhood residents with a bachelor’s degree or higher was associated with increased odds of reversion to MCI in both model configurations (OR = 1.13; 95% CI: 1.02–1.24) (**Fig. 3B**, **3D**).

The final logistic regression model incorporated all ICD code-blocks recorded prior to index dementia diagnosis. Using group-LASSO on the training set, we selected 116 predictors, which were then fit in a standard logistic regression (LOGIT) to estimate coefficients and assess generalizability in the test set. In the held-out test set, 12 predictors were significantly associated with increased or decreased odds of reversion to MCI (**Fig. 4**). Several diagnoses reflecting vascular or systemic conditions and healthcare utilization were associated with increased odds of reversion, including atherosclerosis (OR=2.61, 95% CI: 1.26 – 5.40), general symptoms such as fever (OR=1.60, 95% CI: 1.01 – 2.54), mobility issues (OR=1.79, 95% CI: 1.07 – 2.99), and encounters for immunization (OR=2.58, 95% CI: 1.34 – 4.99). Additional conditions associated with higher odds of reversion included unspecified injuries (OR=2.52, 95% CI: 1.20 – 5.29), diverticular disease of the intestine (OR=2.64; 95% CI: 1.31 – 5.31), and other spondylopathies (OR=2.18; 95% CI: 1.07 – 4.42). Other statistically significant predictors identified in the training set, though not statistically significant in the test set, were consistent with known dementia risk factors. These included Type 2 diabetes (OR=0.73, 95% CI: 0.54 – 0.98), other cerebrovascular diseases (OR=0.55, 95% CI: 0.35 – 0.87), lipoprotein metabolism disorders (OR=1.39; 95% CI: 1.07–1.80), unspecified hearing loss (OR=1.74; 95% CI: 1.56 – 2.62), cataracts (OR=1.70; 95% CI: 1.11 – 2.59), and depressive episodes (OR=1.37; 95% CI: 1.02 – 1.84). Additional predictors from the training set included benign prostatic hyperplasia, malaise and fatigue, chronic obstructive pulmonary disease, pain in the throat/chest, and encounters for special examinations. Finally, the direction and magnitude of associations for demographic variables, including older age, non–memory clinic diagnoses, and Spanish-language preference, were consistent with prior models (**Fig. 4**).

**Figure 4:**
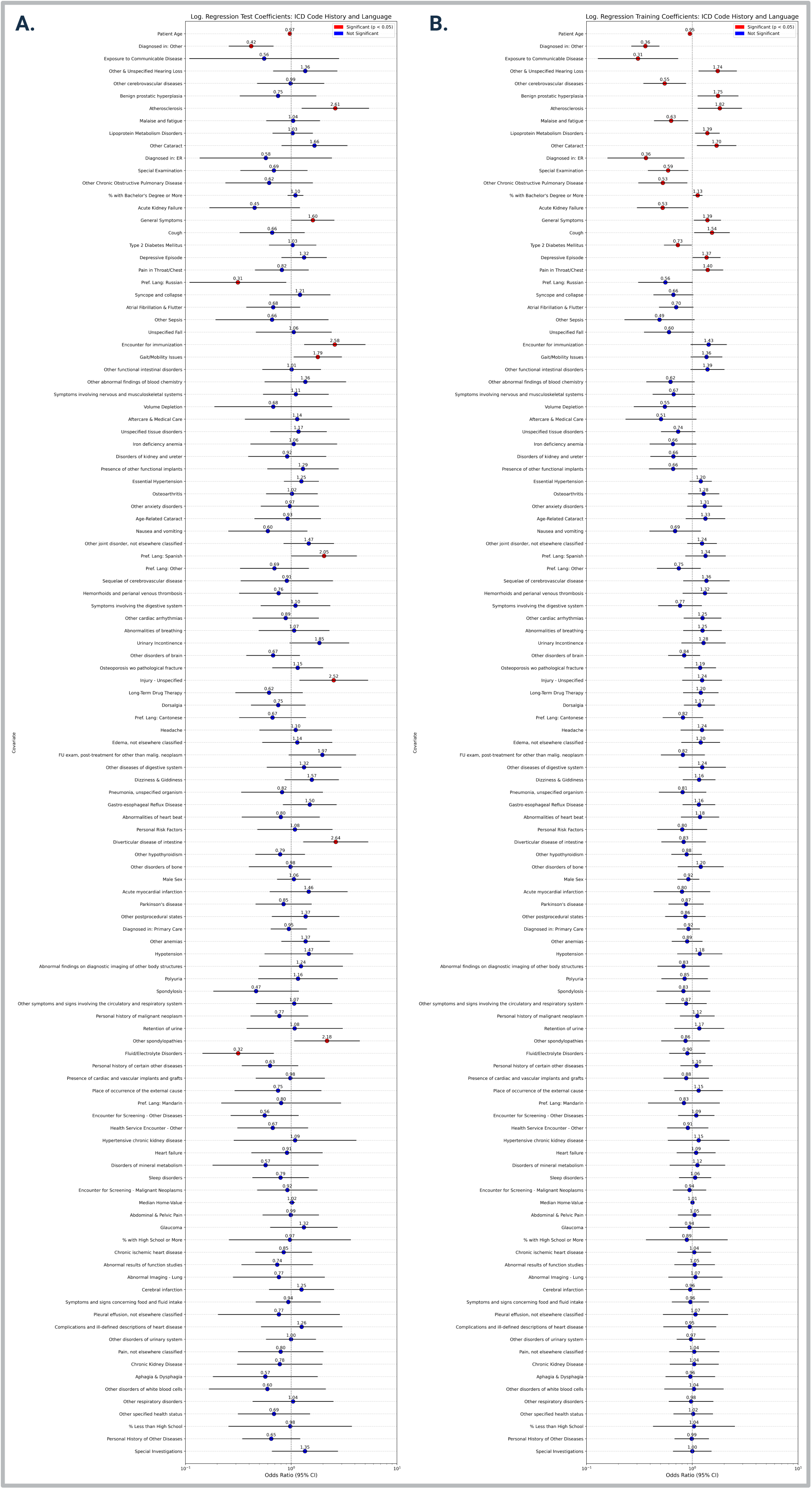
Forest Plots of Logistic Regression (LOGIT) Coefficients from Training and Test Sets for Selected ICD Code Block History Features. **Panels A and B** display coefficients from the test and training sets, respectively. Models were adjusted for patient age at index dementia diagnosis, sex, clinic category, and preferred language. All ICD code blocks shown were selected via Group LASSO feature selection. Red markers indicate statistically significant predictors (p < 0.05); blue markers denote non-significant estimates.

Due to high correlations between race/ethnicity and preferred language, we performed parallel sensitivity analyses for all model configurations in which preferred language was replaced with race/ethnicity. Results of the models adjusting for race/ethnicity instead of preferred language are reported in **Supplemental Figures 1 and 2**.

### 3.3 Random Forest Models Reveal Comorbid Overlap with Conditions Identified in Logistic Regression Models

We trained random forest models using the same covariate configurations applied in the logistic regression analyses. Among the top-ranked features were patient age, clinic type, cardiovascular comorbidities, and ICD block codes for osteoarthritis, depressive episodes, gait and mobility issues, dorsalgia, atrial fibrillation and flutter, and vision problems. Other identified features not overlapping with features identified using Group-LASSO included block-codes for anemias, urinary incontinence, abnormalities of heartbeat, hypothyroidism, disorders of white blood cells, and heart failure. A complete list of important features can be found in **Supplemental Table S9**.

### 3.4 Model Performance: Logistic Regression Outperforms Random Forest While Preserving Interpretability

Model performance was evaluated using AUROC, AUPRC, and threshold-optimized classification metrics on the held-out test set (**Figures 5 – 6; Supplemental Figures 3 – 4; Supplemental Tables S10 – S12**).

**Figure 5:**
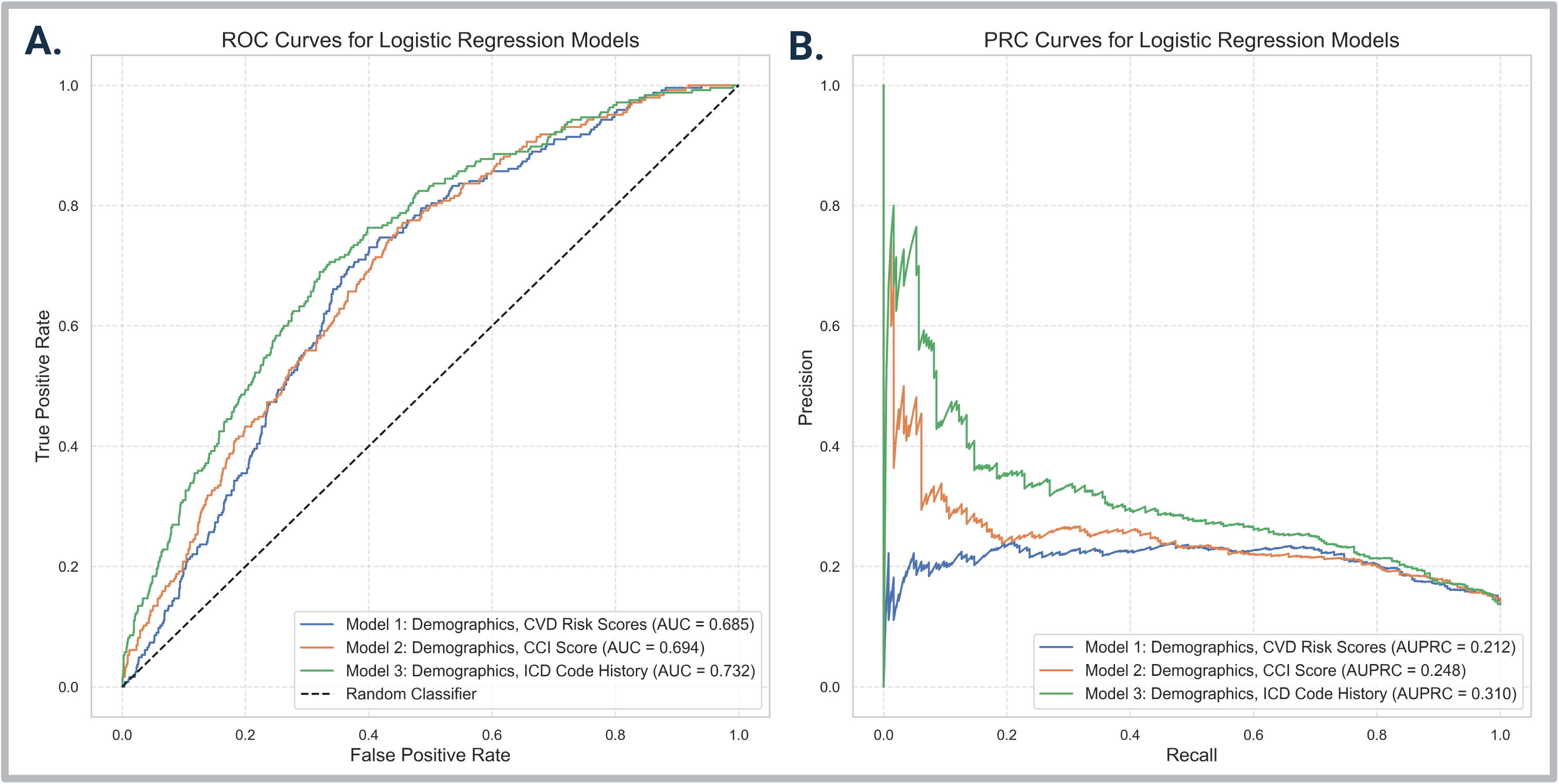
ROC and Precision-Recall Curves for Logistic Regression Models. **A.** Receiver Operating Characteristic (ROC) curves comparing logistic regression (LOGIT) models adjusted for preferred language, across three feature sets, noted in the figure legend. **B.** Precision-Recall Curves (PRC) for the same models, capturing performance in the context of class imbalance. While AUROC captures general model discrimination, AUPRC emphasizes performance of the minority class, patients who reverted to MCI after an index dementia diagnosis.

**Figure 6:**
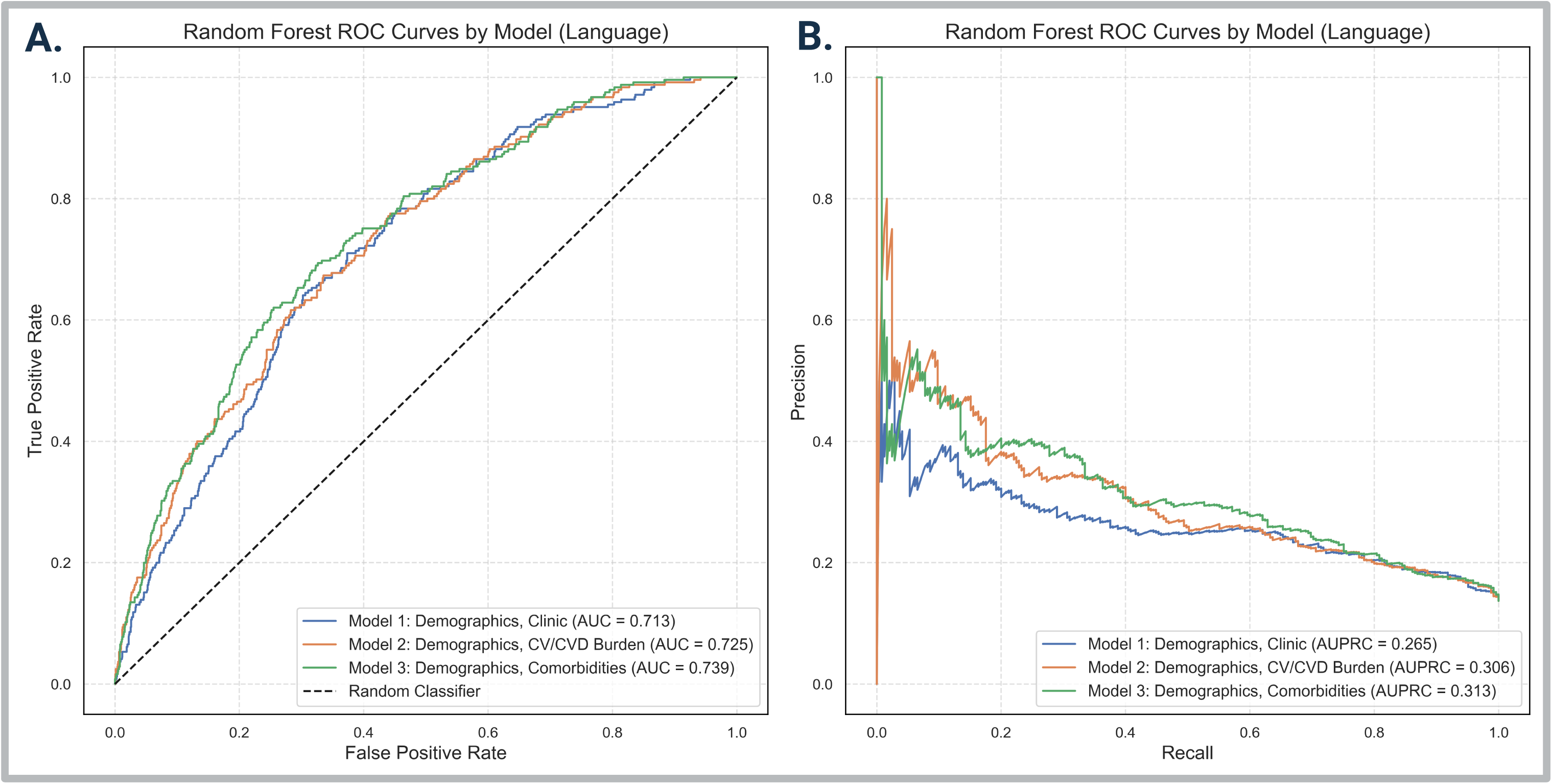
ROC and Precision-Recall Curves for Random Forest Models. **A.** Receiver Operating Characteristic (ROC) curves for random forest models adjusting for preferred language, across three feature sets, depicted in the figure legends. **B.** Precision-Recall Curves (PRC) for the same models, highlighting model performance under class imbalance. As in logistic regression, AUROC captures overall discrimination, while AUPRC better reflects precision and recall for the minority class (patients with diagnostic reversion).

Across all logistic regression models, AUROC ranged from 0.69 to 0.74, and AUPRC ranged from 0.23 to 0.32, with the best-performing model incorporating ICD code-block history, demographics, and clinic type (**Fig. 5**). Threshold optimization using F1 scores on the training set substantially improved sensitivity (TPR range: 0.65 – 0.75) and F1 scores (range: 0.34 – 0.36) compared to the default threshold of 0.5, which originally yielded no positive predictions (**Supplemental Table S11**).

Random forest models exhibited similar performance across matched covariate configurations, with AUROC ranging from 0.72 to 0.73 and AUPRC from 0.27 to 0.31 (**Fig. 6**). After threshold optimization, F1 scores improved up to 0.38, with corresponding gains in recall (TPR range: 0.29 – 0.56) and precision (PPV range: 0.26 – 0.39) (**Supplemental Table S12**). The top performing random forest model, using ICD code-block features and adjusted for preferred language, achieved an AUROC 0.73, AUPRC of 0.31, and F1 score of 0.38 after threshold tuning. **Supplemental Figures 3–4** show that performance was consistent when models were adjusted for race/ethnicity, closely mirroring language-adjusted models.

## 4. Discussion

We examined diagnostic reversion—MCI diagnosis after dementia—in a large outpatient sample of older adults within an academic healthcare system. Using a comparative model framework, we identified clinical, demographic, and care-setting factors associated with this pattern, occurring in 13.7% of patients. Reversion may reflect premature dementia diagnosis or MCI misclassification, with implications for diagnostic accuracy, care planning, and patient burden [5].

Analysis of diagnostic trajectories among reverters revealed patterns of diagnostic instability in routine dementia care. While the median time to MCI reversion was approximately nine months, most patients remained with an MCI diagnosis for a relatively short duration (median ∼3 months) before being re-diagnosed with dementia. The fact that nearly three-quarters (73.7%) of reverters were ultimately reassigned to dementia suggests that reversion may often reflect diagnostic hesitation rather than true clinical improvement. The weak but positive correlation between time to MCI reversion and time spent with an MCI diagnosis suggests that patients with more protracted diagnostic shifts may have presented with more ambiguous clinical features. This is further supported by greater healthcare utilization among reverters, potentially reflecting increased diagnostic complexity.

Our results underscore the complex and often uncertain nature of cognitive diagnosis assignment in real-world-settings. Patients who were younger and had a higher number of CV risk factors were more likely to be reassigned a diagnosis of MCI, raising the possibility that their initial dementia diagnosis was premature. In contrast, patients diagnosed in the ER were less likely to revert to MCI. One interpretation is that patients seen in ER settings may have exhibited more definitive symptoms or presented to these clinics in later stages of cognitive decline [40,41]. We also found that higher CVD Burden scores were associated with lowered odds of reversion, suggesting that a more extensive cardiovascular disease profile may reflect true neurodegenerative progression. Conversely, higher Charlson Comorbidity Index (CCI) scores were linked to increased odds of MCI reversion, possibly reflecting diagnostic overshadowing by competing health conditions. Together, these findings point to meaningful variation in diagnostic certainty based on patient complexity and care setting.

Patients’ preferred language was associated with the likelihood of diagnostic reversion. Spanish-speaking patients were more likely to revert to MCI after an initial dementia diagnosis, raising concerns about potential language barriers in symptom communication and gaps in language-concordant diagnostic care, issues previously documented in neurological health disparities research [42]. This discrepancy may reflect differences in clinical presentation, healthcare-seeking behavior, or provider-related factors. Notably, our study did not capture information on provider language proficiency or cultural concordance between patients and providers, which may influence communication quality and diagnostic certainty [43,44]. Conversely, Russian-speaking patients were significantly *less* likely to experience diagnostic reversion, possibly reflecting later-stage clinical presentation, greater reliance on interpreter-supported care, or the presence of caregivers influencing diagnostic communication and decision-making.

Educational attainment, measured at the census tract level, was associated with diagnostic reversion only in the training set, suggesting limited generalizability. Patients residing in areas with a higher proportion of adults holding a bachelor’s degree or higher had greater odds of reversion, which may reflect increased access to follow-up care or diagnostic reevaluation in socioeconomically advantaged neighborhoods [45–47]. This trend may also align with the demographic context of San Francisco, where 60.4% of adults aged 25+ hold a bachelor’s degree or higher [48]. Median home value was not significantly associated with reversion, potentially due to the overall affluence and limited variation in housing-based socioeconomic factors within our cohort.

We used complementary data-driven approaches to identify diagnostic patterns linked to reversion. Chi-square tests comparing ICD block prevalence between patient groups revealed that patients who reverted to MCI were more likely to present with non-specific or functional symptoms such as dizziness and giddiness, abnormal gait and mobility, depressive episodes, and osteoarthritis (**Figure 2C**). These conditions, while disruptive, may reflect ambiguous clinical presentations or comorbidities that complicate diagnosis, rather than progressive cognitive decline. Conversely, chronic illnesses such as hypertensive kidney disease and heart failure, were more common among patients who remained diagnosed with dementia, supporting the idea that diagnostic stability is more likely when patients present with clear signs of advanced systemic decline.

In our multivariable models adjusting for demographic and clinical covariates, diagnostic reversion was associated with ICD code blocks suggestive of either vague symptoms or ongoing care engagement. In the test set, conditions such as atherosclerosis, immunization encounters, gait abnormalities, unspecified injuries, and diverticular disease were linked to higher odds of reversion. Similar patterns emerged in the training set, including associations with hearing loss, lipid disorders, depression, and cataracts. Several of these conditions are known dementia risk factors [49], suggesting that reversion may reflect diagnostic uncertainty in patients with evolving clinical presentations. In contrast, lower odds of reversion were observed among patients with chronic illnesses (e.g., cerebrovascular disease, chronic obstructive pulmonary disease, acute kidney failure, Type 2 diabetes), consistent with chi-square results and reinforcing the idea that chronic disease burden may anchor diagnostic certainty among patients in more advanced stages of cognitive decline [50]. Together, these results suggest that diagnostic reversion is more likely to occur in patients with non-specific symptoms, or continued clinical monitoring, while stability in dementia diagnosis tends to co-occur with chronic or end stage comorbidities, potentially reflecting clearer underlying pathology.

From a methodological standpoint, logistic regression models slightly outperformed random forests in AUROC (up to 0.74 vs. 0.73) (**Fig. 4 – 5**), while also offering greater interpretability, particularly through group LASSO feature selection, which clarified key predictors such as preferred language and chronic conditions (**Fig. 4 — 6**). Although AUPRC scores were modest across models due to class imbalance, threshold optimization markedly improved sensitivity and precision. At the default probability threshold (0.5), models failed to identify reverters, but tuning for F1 score raised recall to 0.75t in the best logistic regression model. These findings suggest interpretable models, when properly optimized, can match or exceed black-box alternatives in both performance and clinical relevance.

Our findings underscore the clinical and structural complexity of real-world dementia diagnosis. We identified several factors associated with diagnostic reversion, including demographic, clinical and care-setting variables, suggesting meaningful variation in diagnostic certainty and access to follow-up evaluation. These results emphasize the need for more standardized diagnostic pathways, improved integration of longitudinal clinical data, and equitable access to comprehensive evaluations, particularly for population facing linguistic or socioeconomic barriers.

This study has several limitations. First, diagnoses were identified from EHR fields originally intended for billing, which may result in missingness or inconsistent documentation. We did not include unstructured clinical notes or formal cognitive assessments, limiting our ability to determine whether diagnostic reversion reflected clinical improvement or labeling uncertainty. Additionally, we did not include data on pharmacologic treatments that may influence clinical status or diagnostic decision making.

Socioeconomic variables, such as educational attainment and median home value, were measured at the census tract level due to inconsistent or missing individual-level data. These proxies reflect neighborhood context but do not capture individual cognitive reserve or personal education histories. We also lacked information on family support networks, particularly the educational level or advocacy role of adult children, which may shape healthcare navigation and access to diagnostic reevaluation [51–53].

Our outcome of interest – diagnostic reversion– was observed in 13.7% of patients, which may have limited statistical power in test set evaluations. While many associations held consistent directionality, some did not replicate with overlapping confidence intervals. Finally, this study was conducted within a single academic health system (UCSF), which may limit generalizability. UCSF serves a diverse, urban population, but as a tertiary referral center, many patients may have received only specialty care, limiting capture of longitudinal diagnostic histories.

Despite these limitations, our results provide important insights into patterns of diagnostic switching and the contexts in which they arise. Diagnostic reversion may serve as a proxy for clinical uncertainty, prompting closer attention to how diagnoses are assigned, revisited, and communicated across settings. Future research should build on these findings using multisite data, clinical notes, and validated cognitive assessments to better understand diagnostic discrepancies, care pathways, and opportunities for intervention in dementia care.

## Data Availability

All data used and produced in the present study is not publicly available due to HIPAA regulations and exclusive use to trained researchers within the University of California, San Francisco with access to de-identified EHR data.

## Consent Statement

This study was approved by the UCSF Institutional Review Board (IRB #20-32422). Informed consent was not required, as the research involved secondary analysis of electronic health records without direct patient contact, and met criteria for a waiver of consent under applicable regulations.

