## Supplementary figures and images for "Diagnostic Reversion in Dementia Care: A Real-World Analysis of Mild Cognitive Impairment Diagnoses Following Dementia in a Large Electronic Medical Record System"

### Supplemental Figure 1

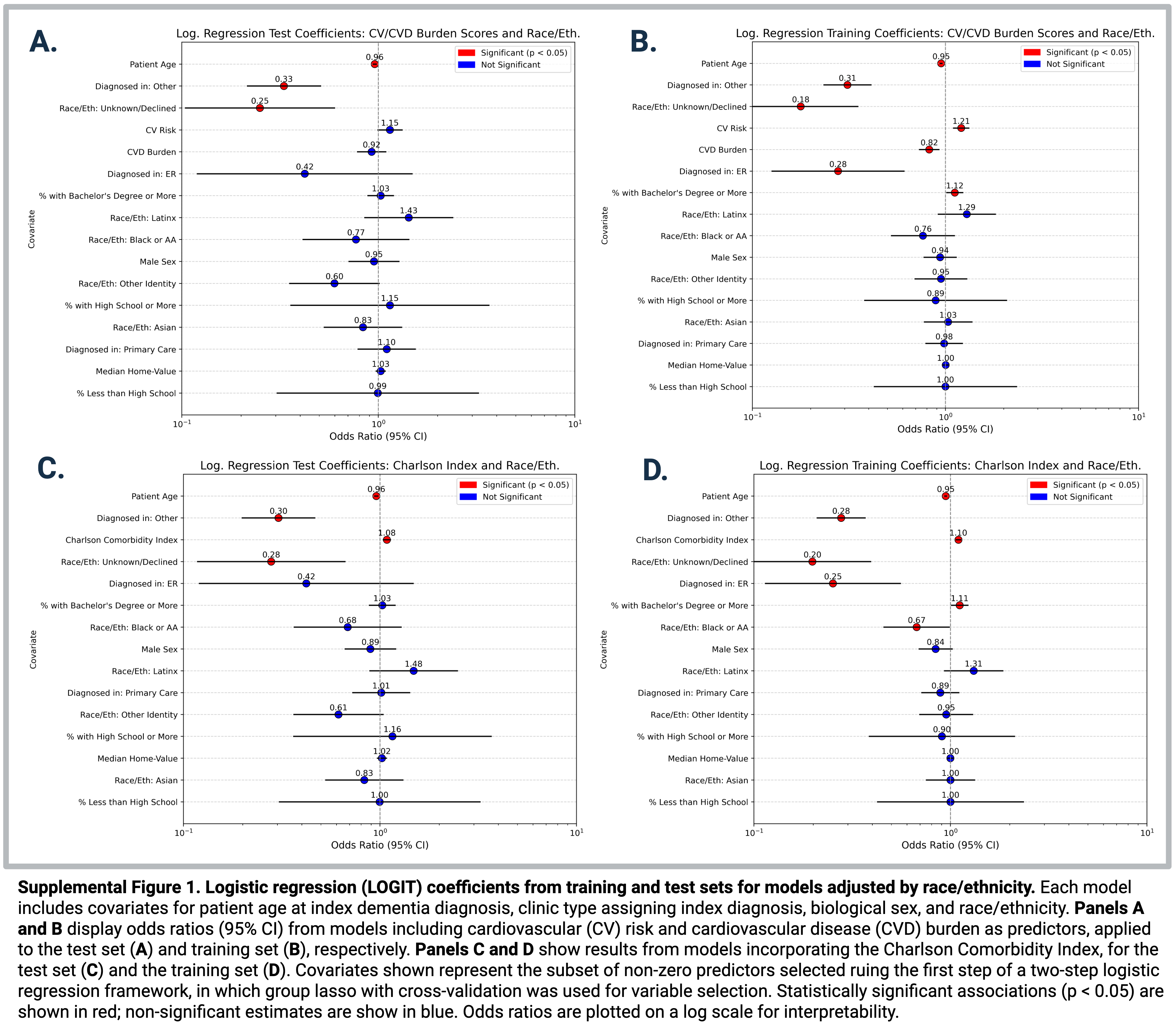

### Supplemental Figure 2

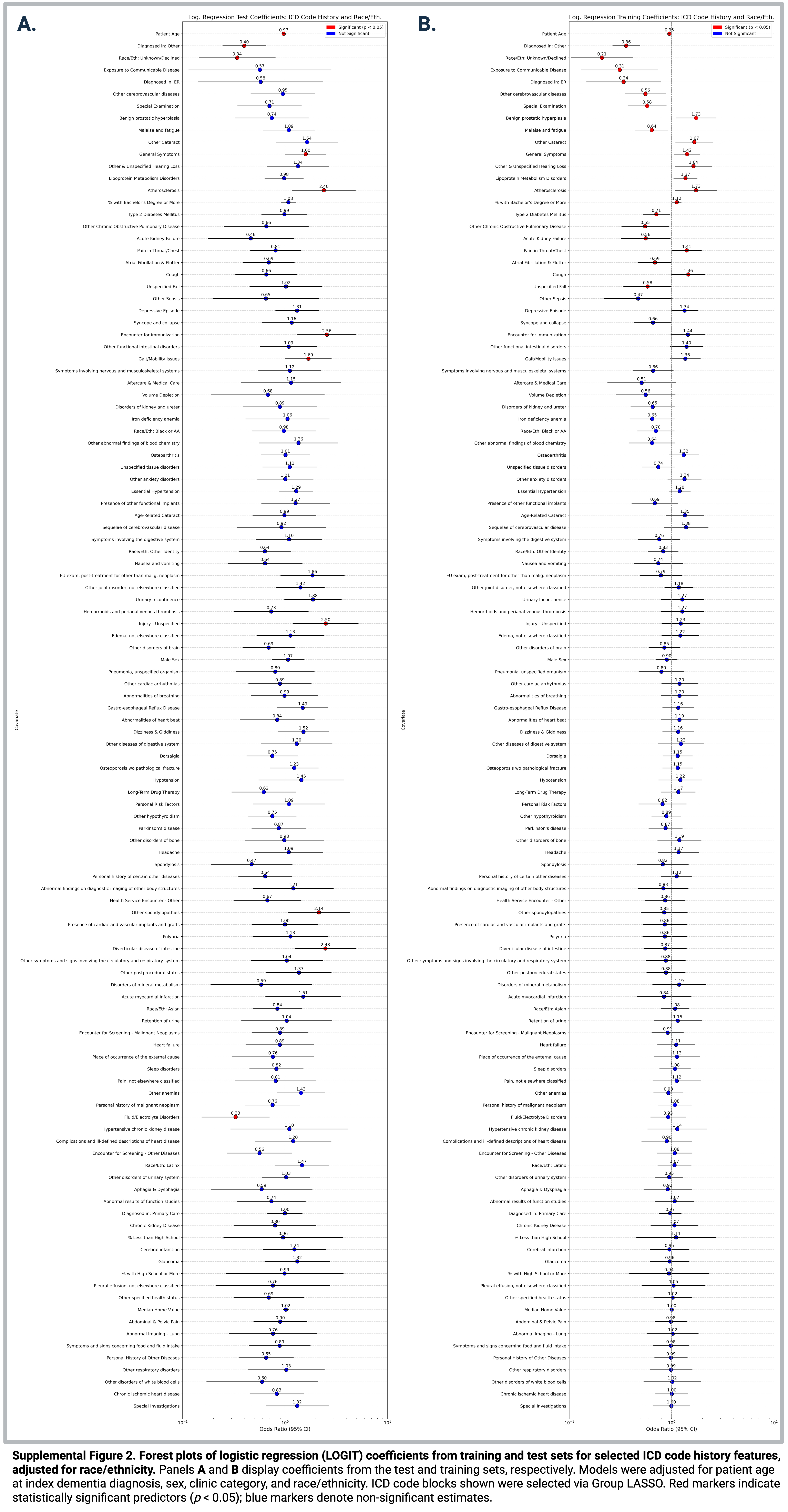

### Supplemental Figure 3

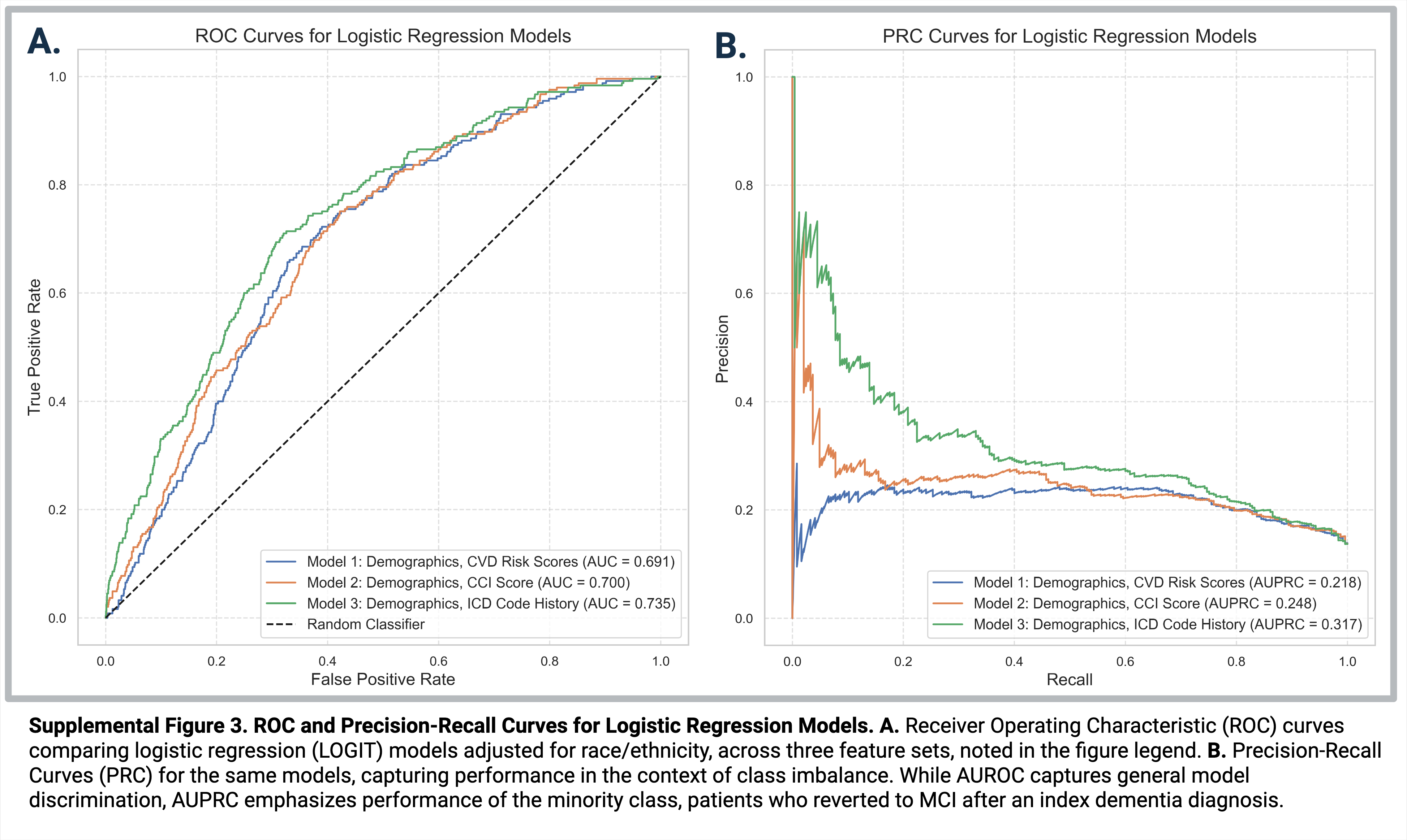

### Supplemental Figure 4

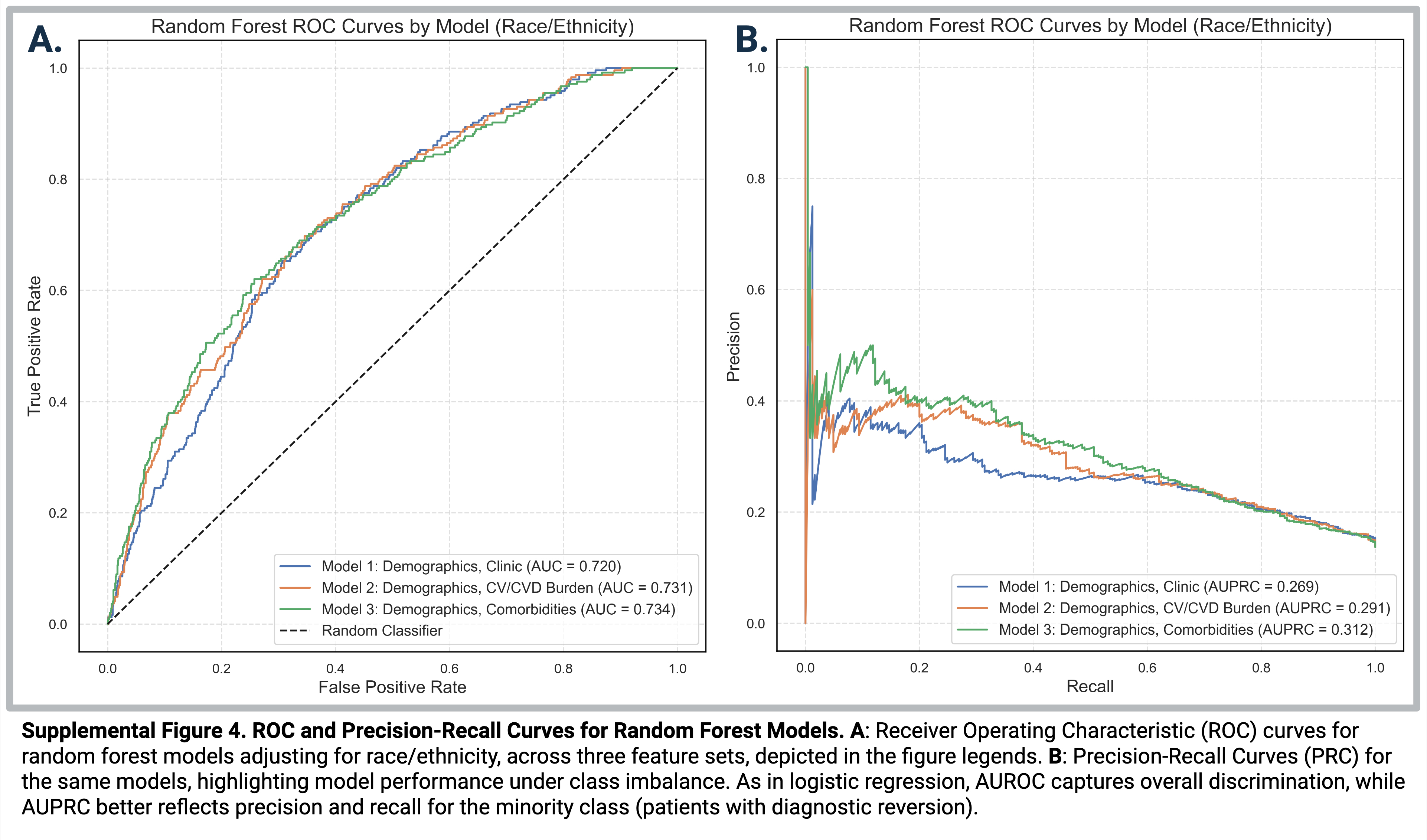
