## Supplemental Tables for "Diagnostic Reversion in Dementia Care: A Real-World Analysis of Mild Cognitive Impairment Diagnoses Following Dementia in a Large Electronic Medical Record System"

**Supplemental Table S1:** ICD Codes Used to Identify MCI Diagnoses

| **ICD Code** | **Description** |
| --- | --- |
| G31.84 | Mild cognitive impairment of uncertain or unknown etiology |
| R41.1 | Anterograde amnesia |
| R41.2 | Retrograde amnesia |
| R41.3 | Other amnesia |
| R41.81 | Age-related cognitive decline |
| R41.82 | Altered mental status, unspecified |
| R41.840 | Attention and concentration deficit |
| R41.841 | Cognitive communication deficit |
| R41.842 | Visuospatial deficit |
| R41.843 | Psychomotor deficit |
| R41.844 | Frontal lobe and executive function deficit |
| R41.89 | Other symptoms and signs involving cognitive functions and awareness |
| R41.9 | Unspecified symptoms and signs involving cognitive functions and awareness |
| F06.8 | Other specified mental disorders due to known physiological condition |
| 331.83 | Mild Cognitive Impairment of uncertain or unknown etiology |
| 780.93 | Memory Loss |
| 797 | Senility without mention of psychosis |
| 780.97 | Altered mental status |
| 799.51 | Attention and concentration deficit |
| 799.52 | Unspecified signs and symptoms involving cognition |
| 799.53 | Visuospatial deficit |
| 799.54 | Psychomotor deficit |
| 799.55 | Frontal lobe and executive function deficit |
| 799.59 | Other signs and symptoms involving cognition |
| 294.8 | Other persistent mental disorders due to conditions classified elsewhere |

**Supplemental Table S2:** ICD Codes Used to Identify Dementia Diagnoses

| **ICD Code** | **Description** |
| --- | --- |
| F00.0 | Dementia in Alzheimer disease |
| F00.1 | Dementia in Alzheimer disease with late onset |
| F00.2 | Dementia in Alzheimer's disease, atypical or mixed type |
| F00.9 | Dementia in Alzheimer's disease, unspecified |
| F01 | Vascular Dementia |
| F01.5 | Vascular Dementia, unspecified severity |
| F01.50 | Vascular Dementia with behavioral disturbance, psychotic disturbance, mood disturbance, and anxiety |
| F01.51 | Vascular Dementia, unspecified severity, with unspecified severity, with agitation |
| F01.518 | Vascular dementia, unspecified severity, with other behavioral disturbance. |
| F01.52 | Vascular Dementia with psychotic disturbance |
| F01.53 | Vascular Dementia with mood disturbance |
| F01.54 | Vascular Dementia with Anxiety |
| F01.A | Vascular Dementia, mild |
| F01.A0 | Vascular Dementia mild, without behavioral disturbance, psychotic disturbance, mood disturbance and anxiety |
| F01.A1 | Vascular Dementia, mild, with behavioral disturbance |
| F01.A11 | Vascular Dementia, mild, with agitation |
| F01.A18 | Vascular dementia, mild, with other behavioral disturbance |
| F01.A2 | Vascular dementia with psychotic disturbance |
| F01.A3 | Vascular dementia with mood disturbance |
| F01.A4 | Vascular Dementia with anxiety |
| F01.B | Vascular Dementia, moderate |
| F01.B0 | Vascular Dementia, moderate, without behavioral disturbance, psychotic disturbance, mood disturbance and anxiety |
| F01.B1 | Vascular Dementia, moderate, with behavioral disturbance |
| F01.B11 | Vascular Dementia, moderate, with agitation |
| F01.B18 | Vascular Dementia, moderate, with other behavioral disturbance |
| F01.B2 | Vascular dementia, moderate, with psychotic disturbance |
| F01.B3 | Vascular Dementia, moderate, with mood disturbance |
| F01.B4 | Vascular Dementia, moderate, with anxiety |
| F01.C | Vascular Dementia, severe |
| F01.C0 | Vascular Dementia, severe, without behavioral disturbance, psychotic disturbance, mood disturbance, and anxiety |
| F01.C1 | Vascular Dementia, severe, with behavioral disturbance |
| F01.C11 | Vascular Dementia, severe, with agitation |
| F01.C18 | Vascular Dementia, severe, with other behavioral disturbance |
| F01.C2 | Vascular Dementia, severe, with psychotic disturbance |
| F01.C3 | Vascular Dementia, severe, with mood disturbance |
| F01.C4 | Vascular Dementia, severe, with anxiety |
| F01.2 | Subcortical Vascular Dementia |
| F01.3 | Mixed Cortical and subcortical vascular dementia |
| F01.8 | Other Vascular Dementia |
| F02 | Dementia in other diseases classified elsewhere |
| F02.8 | Dementia in other diseases classified elsewhere, unspecified severity |
| F02.80 | Dementia in other diseases classified elsewhere, without behavioral disturbance, psychotic disturbance, mood disturbance, and anxiety |
| F02.81 | Dementia in other diseases classified elsewhere, unspecified severity with behavioral disturbance |
| F02.811 | Dementia in other disease classified elsewhere, unspecified severity, with agitation |
| F02.818 | Dementia in other disease classified elsewhere, unspecified severity, with other behavioral disturbance |
| F02.82 | Dementia in other diseases classified elsewhere with psychotic disturbance |
| F02.83 | Dementia in other diseases classified elsewhere with mood disturbance |
| F02.84 | Dementia in other diseases classified elsewhere with anxiety |
| F02.A | Dementia in other diseases classified elsewhere, mild |
| F02.A0 | Dementia in other diseases classified elsewhere, mild, without behavioral disturbance, psychotic disturbance, mood disturbance, and anxiety |
| F02.A1 | Dementia in other diseases classified elsewhere, mild, with behavioral disturbance |
| F02.A11 | Dementia in other diseases classified elsewhere, mild, with agitation |
| F02.A18 | Dementia in other diseases classified elsewhere, mild, with other behavioral disturbance |
| F02.A2 | Dementia in other diseases classified elsewhere, mild, with psychotic disturbance |
| F02.A3 | Dementia in other diseases classified elsewhere, mild, with mood disturbance |
| F02.A4 | Dementia in other diseases classified elsewhere, mild, with anxiety |
| F02.B | Dementia in other diseases classified elsewhere, moderate |
| F02.B0 | Dementia in other diseases classified elsewhere, moderate, without behavioral disturbance, mood disturbance, and anxiety |
| F02.B1 | Dementia in other diseases classified elsewhere, moderate, with behavioral disturbance |
| F02.B11 | Dementia in other diseases classified elsewhere, moderate, with agitation |
| F02.B18 | Dementia in other diseases classified elsewhere, moderate, with other behavioral disturbance |
| F02.B2 | Dementia in other diseases classified elsewhere, moderate, with psychotic disturbance |
| F02.B3 | Dementia in other diseases classified elsewhere, moderate, with mood disturbance |
| F02.B4 | Dementia in other diseases classified elsewhere, moderate, with anxiety |
| F02.C | Dementia in other diseases classified elsewhere, severe |
| F02.C0 | Dementia in other diseases classified elsewhere, severe, without behavioral disturbance, psychotic disturbance, mood disturbance, and anxiety |
| F02.C1 | Dementia in other diseases classified elsewhere, severe, with behavioral disturbance |
| F02.C11 | Dementia in other diseases classified elsewhere, severe, with agitation |
| F02.C18 | Dementia in other diseases classified elsewhere, severe, with other behavioral disturbance. |
| F02.C2 | Dementia in other diseases classified elsewhere, severe, with psychotic disturbance |
| F02.C3 | Dementia in other diseases classified elsewhere, severe, with mood disturbance |
| F02.C4 | Dementia in other diseases classified elsewhere, severe, with anxiety |
| F03 | Unspecified Dementia |
| F03.9 | Unspecified dementia, unspecified severity |
| F03.90 | Unspecified dementia, unspecified severity without behavioral disturbance, psychotic disturbance, mood disturbance, and anxiety |
| F03.91 | Unspecified dementia, unspecified severity, with behavioral disturbance |
| F03.911 | Unspecified dementia, unspecified severity, with agitation |
| F03.918 | Unspecified dementia, unspecified severity, with other behavioral disturbance. |
| F03.92 | Unspecified dementia, unspecified severity with psychotic disturbance |
| F03.93 | Unspecified dementia, unspecified severity with mood disturbance |
| F03.94 | Unspecified dementia, unspecified severity with anxiety |
| F03.A | Unspecified dementia, mild |
| F03.A0 | Unspecified dementia, mild without behavioral disturbance, psychotic disturbance, mood disturbance, and anxiety. |
| F03.A1 | Unspecified dementia, mild, with behavioral disturbance |
| F03.A11 | Unspecified dementia, mild, with agitation |
| F03.A18 | Unspecified dementia, mild, with other behavioral disturbance |
| F03.A2 | Unspecified dementia, mild, with psychotic disturbance |
| F03.A3 | Unspecified dementia, mild, with mood disturbance |
| F03.A4 | Unspecified dementia, mild, with anxiety |
| F03.B | Unspecified dementia, moderate |
| F03.B0 | Unspecified dementia, moderate, without behavioral disturbance, psychotic disturbance, mood disturbance, and anxiety |
| F03.B1 | Unspecified dementia, moderate, with behavioral disturbance |
| F03.B11 | Unspecified dementia, moderate, with agitation |
| F03.B18 | Unspecified dementia, moderate, with other behavioral disturbance |
| F03.B2 | Unspecified dementia, moderate, with psychotic disturbance |
| F03.B3 | Unspecified dementia, moderate, with mood disturbance |
| F03.B4 | Unspecified dementia, moderate, with anxiety |
| F03.C | Unspecified dementia, severe |
| F03.C0 | Unspecified dementia, severe, without behavioral disturbance, psychotic disturbance, mood disturbance, and anxiety |
| F03.C1 | Unspecified dementia, severe, with behavioral disturbance |
| F03.C11 | Unspecified dementia, severe, with agitation |
| F03.C18 | Unspecified dementia, severe, with other behavioral disturbance |
| F03.C2 | Unspecified dementia, severe, with psychotic disturbance |
| F03.C3 | Unspecified dementia, severe, with mood disturbance |
| F03.C4 | Unspecified dementia, severe, with anxeity |
| G30 | Alzheimer's Disease |
| G30.0 | Alzheimer's Disease with early onset |
| G30.1 | Alzheimer's Disease with late onset |
| G30.8 | Other Alzheimer's Disease |
| G30.9 | Alzheimer's Disease, unspecified |
| G31.0 | Frontotemporal Dementia |
| G31.01 | Pick's Disease |
| G31.1 | Senile degeneration of brain, not elsewhere classified |
| G31.09 | Other Frontotemporal neurocognitive disorder |
| G31.83 | Neurocognitive Disorder with Lewy bodies |
| 290 . 0 | Senile dementia, simple type |
| 290.1 | Presenile dementia |
| 290.2 | Senile dementia, depressed or paranoid type |
| 290.3 | Unspecified dementia, unspecified severity, without behavioral disturbance, psychotic disturbance, mood disturbance, and anxiety |
| 290.4 | Vascular dementia, unspecified severity, without behavioral disturbance |
| 290.4 0 | Vascular dementia, uncomplicated |
| 290.41 | Vascular dementia, unspecified severity, with behavioral disturbance |
| 290.9 | Unspecified dementia, unspecified severity, without behavioral disturbance, psychotic disturbance, mood disturbance, and anxiety |
| 294.1 0 | Dementia in other diseases classified elsewhere, unspecified severity, without behavioral disturbance, psychotic disturbance, mood disturbance, and anxiety |
| 294.11 | Dementia in conditions classified elsewhere with behavioral disturbance |
| 294.2 | Dementia with no underlying cause or specified cause. |
| 294.2 0 | Unspecified dementia, unspecified severity, without behavioral disturbance, psychotic disturbance, mood disturbance, and anxiety |
| 294.21 | Dementia with unspecified severity and behavioral disturbance |
| 331. 0 | Alzheimer's Disease |
| 331.2 | Senile degeneration of the brain |
| 331.8 | Cerebral Degeneration |
| 331.19 | Other frontotemporal neurocognitive disorder |
| 331.82 | Neurocognitive disorder with Lewy bodies |

**Supplemental Table S3:** Generalized Categories for UCSF Clinics Labeled as “Other”

| **Clinic Category** | **Number of Clinics** |
| --- | --- |
| Uncategorized | 180 |
| Surgery (Non-Oncology) | 31 |
| Cardiology | 20 |
| Oncology – Medical | 20 |
| Endocrinology / Diabetes | 9 |
| Neurology / ALS / EEG | 9 |
| Oncology – Surgical | 9 |
| Laboratory / Pathology | 7 |
| Dermatology | 6 |
| Pediatrics / NICU | 3 |
| Audiology / Speech | 1 |
| Behavioral Health | 1 |
| Clinical Research / Trials | 1 |

**Supplemental Table S4:** Cardiovascular Diseases (CVD Burden) ICD Block Codes

| **ICD Code** | **Description** |
| --- | --- |
| I50 | Heart failure |
| I51 | Complications and ill-defined descriptions of heart disease |
| I48 | Atrial fibrillation and flutter |
| Z95 | Presence of cardiac and vascular implants and grafts |
| Z98 | Other postprocedural states |
| I21 | Acute myocardial infarction |
| I46 | Cardiac arrest |
| I52 | Other heart disorders in diseases classified elsewhere |

**Supplemental Table S5:** Cardiovascular Risk Burden (CV Burden) ICD Block Codes

| **ICD Code** | **Description** |
| --- | --- |
| I10 | Essential (primary) hypertension |
| E11 | Type 2 diabetes mellitus |
| E78 | Disorders of lipoprotein metabolism and other lipidemias |
| R73 | Elevated blood glucose level |
| I73 | Other peripheral vascular diseases |

**Supplemental Table S6:** Healthcare Utilization Metrics by Patient Group

| **Healthcare Utilization**  **Metric** | **Comparison Group**  **(Reverters)**  **Median**  **[IQR]** | **Reference Group**  **(Dementia Only) Median**  **[IQR]** | **p-value** |
| --- | --- | --- | --- |
| **Total Days in EHR System** | 3,076  [1,570–5,461] | 1,481  [175–4,463] | < 0.05 |
| **# of Diagnoses** | 127  [36–317] | 46  [13–138] | < 0.05 |
| **# of Unique Conditions** | 42  [16–80] | 19  [7–42] | < 0.05 |
| **# of EHR Visits** | 43  [14–101] | 13  [4–39] | < 0.05 |
| **# of Days in EHR System Before Index Dementia** | 1,128  [152–2,736] | 1,176  [43–4,198] | 0.51 |
| **# of Diagnoses Before Index Dementia** | 23  [5–91] | 23  [5–88] | 0.95 |
| **# of Unique Conditions Before Index Dementia** | 13  [4–33] | 12  [4–31] | 0.53 |
| **# of EHR Visits Before Index Dementia** | 9  [3–32] | 8  [2–28] | 0.03 |

**Supplemental Table S7:** Proportion of Patients with UCSF EHR Data Prior to Index

| **Patient Group** | **# of Patients with EHR Data Before Index Dementia Diagnosis** | **Total Patients** | **Percent**  **(%)** |
| --- | --- | --- | --- |
| **Dementia Only** | 3790 | 5148 | 73.6 |
| **Reverter** | 669 | 818 | 81.8 |

**Supplemental Table S8:** Statistically Significant ICD Block Codes Identified via Chi-Square Analyses.

| **ICD Block Code** | **ICD Code Description** |
| --- | --- |
| 780 | General symptoms |
| N17 | Acute kidney failure |
| H26 | Other cataract |
| A41 | Other sepsis |
| E87 | Other disorders of fluid, electrolyte and acid-base balance |
| H25 | Age-related cataract |
| R91 | Abnormal findings on diagnostic imaging of lung |
| Z23 | Encounter for immunization |
| Z20 | Suspected exp. to communicable diseases |
| R42 | Dizziness and giddiness |
| Z79 | Long term (current) drug therapy |
| N18 | Chronic kidney disease (CKD) |
| E86 | Volume depletion |
| M19 | Other and unspecified osteoarthritis |
| Z51 | Encounter for other aftercare and medical care |
| G93 | Other disorders of brain |
| J90 | Pleural effusion, not elsewhere classified |
| W19 | Unspecified fall |
| D72 | Other disorders of white blood cells |
| I48 | Atrial fibrillation and flutter |
| T14 | Injury of unspecified body region |
| R94 | Abnormal results of function studies |
| Z01 | Encounter for other special examination without complaint, suspected or reported diagnosis |
| R26 | Abnormalities of gait and mobility |
| R13 | Aphagia and dysphagia |
| M54 | Dorsalgia |
| H91 | Other and unspecified hearing loss |
| I12 | Hypertensive chronic kidney disease |
| Z12 | Encounter for screening of malignant neoplasms |
| F32 | Depressive Episode |
| H40 | Glaucoma |
| K57 | Diverticular disease of intestine |
| Z86 | Personal history of certain other diseases |
| M81 | Osteoporosis without current pathological |
| K64 | Hemorrhoids and perianal venous thombosis |
| R29 | Other symptoms and signs involving the nervous system |
| V72 | Special investigations and examinations |
| J98 | Other respiratory disorders |
| J18 | Pneumonia, unspecified organism |

**Supplemental Table S9:** Important Features from Random Forest Models

| **Feature** | **RF Importance (%)** |
| --- | --- |
| Patient Age | 12.64 |
| Diagnosed in: Other | 8.5 |
| General Symptoms | 7.26 |
| % with Bachelor's Degree or More | 2.92 |
| Median Home-Value | 2.54 |
| Age-Related Cataract | 2.52 |
| Acute Kidney Failure | 2.43 |
| Other Cataract | 2.33 |
| Encounter for immunization | 1.99 |
| Dizziness & Giddiness | 1.72 |
| Fluid/Electrolyte Disorders | 1.5 |
| Other Sepsis | 1.43 |
| Osteoarthritis | 1.33 |
| Diagnosed in: Primary Care | 1.28 |
| Depressive Episode | 1.16 |
| Other & Unspecified Hearing Loss | 1.1 |
| Cough | 1.09 |
| Long-Term Drug Therapy | 1.09 |
| % with High School or More | 1.0 |
| Injury - Unspecified | 0.97 |
| Unspecified Fall | 0.96 |
| Special Examination | 0.94 |
| Abnormal Imaging - Lung | 0.92 |
| % Less than High School | 0.91 |
| Gait/Mobility Issues | 0.91 |
| Dorsalgia | 0.86 |
| Exposure to Communicable Disease | 0.83 |
| Lipoprotein Metabolism Disorders | 0.77 |
| Personal History of Other Diseases | 0.76 |
| Symptoms involving nervous and musculoskeletal systems | 0.75 |
| Other anxiety disorders | 0.74 |
| Personal history of certain other diseases | 0.72 |
| Other disorders of brain | 0.71 |
| Encounter for Screening - Other Diseases | 0.71 |
| Abnormal results of function studies | 0.69 |
| Atrial Fibrillation & Flutter | 0.67 |
| Encounter for Screening - Malignant Neoplasms | 0.66 |
| Pref. Lang: Spanish | 0.65 |
| Pain in Throat/Chest | 0.64 |
| Hemorrhoids and perianal venous thrombosis | 0.64 |
| Glaucoma | 0.63 |
| Aftercare & Medical Care | 0.62 |
| Malaise and fatigue | 0.6 |
| Other functional intestinal disorders | 0.6 |
| Other respiratory disorders | 0.59 |
| Volume Depletion | 0.55 |
| Abdominal & Pelvic Pain | 0.55 |
| Chronic Kidney Disease | 0.54 |
| Other joint disorder, not elsewhere classified | 0.53 |
| Disorders of kidney and ureter | 0.52 |
| Headache | 0.52 |
| Other disorders of urinary system | 0.52 |
| Pref. Lang: Other | 0.51 |
| Gastro-esophageal Reflux Disease | 0.51 |
| Pleural effusion, not elsewhere classified | 0.5 |
| Heart failure | 0.49 |
| Other cardiac arrhythmias | 0.49 |
| Health Service Encounter - Other | 0.48 |
| Other disorders of white blood cells | 0.48 |
| Unspecified tissue disorders | 0.47 |
| Other hypothyroidism | 0.46 |
| Other abnormal findings of blood chemistry | 0.45 |
| Special Investigations | 0.43 |
| Other anemias | 0.43 |
| Osteoporosis wo pathological fracture | 0.42 |
| Abnormal findings on diagnostic imaging of other body structures | 0.41 |
| Other symptoms and signs involving the circulatory and respiratory system | 0.41 |
| Chronic ischemic heart disease | 0.4 |
| Personal Risk Factors | 0.4 |
| Diverticular disease of intestine | 0.39 |
| Urinary Incontinence | 0.39 |
| Abnormalities of heart beat | 0.38 |
| Type 2 Diabetes Mellitus | 0.38 |
| Male Sex | 0.37 |
| Other cerebrovascular diseases | 0.36 |
| Retention of urine | 0.35 |
| Polyuria | 0.35 |
| Spondylosis | 0.35 |

**Supplemental Table S10:** Model Descriptions

| **Model** | **Model Description (Covariate Adjustment)** |
| --- | --- |
| 1A | Demographics + Clinic Type + CV/CVD Scores (Pref. Language) |
| 1B | Demographics + Clinic Type + CV/CVD Scores (Race/Ethnicity) |
| 2A | Demographics + Clinic Type + CCI Scores (Pref. Language) |
| 2B | Demographics + Clinic Type + CCI Scores (Race/Ethnicity) |
| 3A | Demographics + Clinic Type + ICD Block Code History (Pref. Language) |
| 3B | Demographics + Clinic Type + ICD Block Code History (Race/Ethnicity) |

**Supplemental Table S11:** Threshold Moving for Class Imbalance in Logistic Regression Models

| **Model** | **Threshold** | **Threshold Value** | **TPR** | **TNR** | **AUROC** | **AUPRC** | **F1** | **Precision** | **Recall** |
| --- | --- | --- | --- | --- | --- | --- | --- | --- | --- |
| **1A** | Default  (0.5) | 0.5 | 0.0 | 1.0 | 0.69 | 0.21 | 0.0 | 0.0 | 0.0 |
|  | Optimized  (F1 on Train) | 0.16 | 0.65 | 0.66 | 0.69 | 0.21 | 0.34 | 0.23 | 0.65 |
| **1B** | Default  (0.5) | 0.5 | 0.0 | 1.0 | 0.69 | 0.22 | 0.0 | 0.0 | 0.0 |
|  | Optimized  (F1 on Train) | 0.17 | 0.63 | 0.68 | 0.69 | 0.22 | 0.35 | 0.24 | 0.63 |
| **2A** | Default  (0.5) | 0.5 | 0.0 | 1.0 | 0.69 | 0.25 | 0.0 | 0.0 | 0.0 |
|  | Optimized  (F1 on Train) | 0.15 | 0.71 | 0.58 | 0.69 | 0.25 | 0.33 | 0.21 | 0.71 |
| **2B** | Default  (0.5) | 0.5 | 0.0 | 1.0 | 0.7 | 0.25 | 0.0 | 0.0 | 0.0 |
|  | Optimized  (F1 on Train) | 0.16 | 0.70 | 0.61 | 0.7 | 0.25 | 0.34 | 0.22 | 0.70 |
| **3A** | Default  (0.5) | 0.5 | 0.0 | 1.0 | 0.73 | 0.31 | 0.0 | 0.0 | 0.0 |
|  | Optimized  (F1 on Train) | 0.17 | 0.65 | 0.69 | 0.73 | 0.31 | 0.36 | 0.25 | 0.65 |
| **3B** | Default  (0.5) | 0.5 | 0.0 | 1.0 | 0.74 | 0.32 | 0.0 | 0.0 | 0.0 |
|  | Optimized  (F1 on Train) | 0.15 | 0.75 | 0.61 | 0.74 | 0.32 | 0.36 | 0.23 | 0.75 |

**Supplemental Table S12:** Threshold Moving for Class Imbalance in Random Forest Models

| **Model** | **Threshold** | **Threshold**  **Value** | **TPR** | **TNR** | **AUROC** | **AUPRC** | **F1** | **Precision** | **Recall** |
| --- | --- | --- | --- | --- | --- | --- | --- | --- | --- |
| 1A | Default (0.5) | 0.5 | 0.0 | 1.0 | 0.71 | 0.27 | 0.0 | 0.0 | 0.0 |
|  | Optimized (F1 on Train) | 0.20 | 0.37 | 0.84 | 0.71 | 0.27 | 0.31 | 0.27 | 0.37 |
| 1B | Default (0.5) | 0.5 | 0.0 | 1.0 | 0.72 | 0.27 | 0.0 | 0.0 | 0.0 |
|  | Optimized (F1 on Train) | 0.20 | 0.4 | 0.82 | 0.72 | 0.27 | 0.32 | 0.26 | 0.4 |
| 2A | Default (0.5) | 0.5 | 0.0 | 1.0 | 0.73 | 0.31 | 0.0 | 0.0 | 0.0 |
|  | Optimized (F1 on Train) | 0.19 | 0.4 | 0.86 | 0.73 | 0.31 | 0.35 | 0.31 | 0.4 |
| 2B | Default (0.5) | 0.5 | 0.0 | 1.0 | 0.73 | 0.29 | 0.0 | 0.0 | 0.0 |
|  | Optimized (F1 on Train) | 0.19 | 0.45 | 0.84 | 0.73 | 0.29 | 0.36 | 0.31 | 0.45 |
| 3A | Default (0.5) | 0.5 | 0.0 | 1.0 | 0.73 | 0.31 | 0.0 | 0.0 | 0.0 |
|  | Optimized (F1 on Train) | 0.16 | 0.56 | 0.79 | 0.73 | 0.31 | 0.38 | 0.29 | 0.56 |
| 3B | Default (0.5) | 0.5 | 0.0 | 1.0 | 0.73 | 0.31 | 0.0 | 0.0 | 0.0 |
|  | Optimized (F1 on Train) | 0.23 | 0.29 | 0.93 | 0.73 | 0.31 | 0.34 | 0.39 | 0.29 |
